# Uveitis with teclistamab and elranatamab: infection or inflammation?

**DOI:** 10.64898/2026.09.29.26364283

**Authors:** Yupeng Wang, Yuhao Guo

**Author notes:** Correspondence: Yuhao Guo, MD, Department of Medicine, Danbury Hospital, 24 Hospital Ave, Danbury, CT, USA. These authors contributed equally to this work.

## Abstract

**Background:** In 2026, the US Food and Drug Administration (FDA) identified uveitis as a potential signal of serious risk for the B-cell maturation antigen (BCMA) × CD3 bispecific antibodies teclistamab and elranatamab. Uveitis in this setting may reflect opportunistic infection or immune-mediated inflammation, which require opposite management.

**Methods:** We analyzed FDA Adverse Event Reporting System (FAERS) reports from July 2020 to June 2026 (8,690,601 reports after deduplication) using a case–non-case design, calculating reporting odds ratios (RORs) and the information component for primary-suspect reports. Comparators were belantamab mafodotin, talquetamab and CD3 bispecific antibodies used outside myeloma. Every primary-suspect uveitis report (n = 76) was classified as infectious, probable immune-mediated (hereafter immune-mediated) or indeterminate by a physician blinded to a rule-based algorithm.

**Results:** Uveitis was reported in 18 of 2,995 teclistamab and 29 of 1,691 elranatamab reports (ROR 4.10 [95% CI 2.58–6.52] and 11.85 [8.21–17.12]). Of these 47 cases, 20 were infectious (18 cytomegalovirus chorioretinitis; median onset 92 days), and infection-specified uveitis was disproportionately reported for both agents and for CD3 bispecific antibodies used outside myeloma. Thirteen were immune-mediated, arose early (median onset 4 days) and were co-reported with cytokine release syndrome in six; immune-mediated uveitis showed a signal only for elranatamab (8 cases; ROR 3.59, 1.79–7.18), not for teclistamab or any comparator. Six of the eight elranatamab cases originated from Japan. Agreement between physician and algorithm was 98.7%.

**Conclusions:** The FDA uveitis signal appears to comprise a late infectious uveitis shared by most T-cell–redirecting bispecific antibodies examined and an early immune-mediated uveitis that showed a signal only with elranatamab. These hypothesis-generating findings support prompt ophthalmological assessment of new visual symptoms and exclusion of cytomegalovirus retinitis before corticosteroids are started.

## Introduction

B-cell maturation antigen (BCMA) × CD3 bispecific antibodies, teclistamab and elranatamab, are now widely used for relapsed or refractory multiple myeloma.^1,2^ Uveitis has been described in teclistamab-treated patients,^3^ and in its January–March 2026 update the US Food and Drug Administration (FDA) identified uveitis as a potential signal of serious risk for both agents.^4^ Uveitis in this setting may reflect opportunistic infection or immune-mediated inflammation, which require opposite management. Here, we examined which of these underlies the signal in the FDA Adverse Event Reporting System (FAERS).

## Methods

We analyzed FAERS reports from July 2020 to June 2026 (8,690,601 reports after deduplication; Supplementary Figure S1) using a case–non-case design, calculating reporting odds ratios (RORs) and the information component for primary-suspect reports, each compared with all other reports.^5,6^ A signal was defined as a lower 95% confidence limit of the ROR >1 with at least three cases, confirmed by a lower 95% credibility limit of the information component (IC025) >0. Comparators were belantamab mafodotin, a BCMA-directed antibody–drug conjugate; talquetamab, a GPRC5D × CD3 bispecific antibody used in myeloma;^7^ and CD3 bispecific antibodies used outside myeloma (pooled; Supplementary Table S1). Uveitis reports were split by whether an infection was specified, and each was classified by a physician blinded to a rule-based algorithm as infectious (pathogen-specific terms), probable immune-mediated (hereafter immune-mediated; onset ≤30 days, co-reported cytokine release syndrome or positive dechallenge/rechallenge, without evidence of infection) or indeterminate (agreement 98.7%; Supplementary Tables S1–S3). Because reference reports could not be reviewed individually, all reference reports with a uveitis term not specifying an infection were counted as events for immune-mediated uveitis, a conservative approach that biases the ROR towards the null. Time to onset was calculated from the first dose, excluding reports with missing or implausible dates. Reporting followed the READUS-PV guideline.^8^

## Results

Uveitis was reported in 18 of 2,995 teclistamab and 29 of 1,691 elranatamab reports (ROR 4.10 [95% CI 2.58–6.52] and 11.85 [8.21–17.12]; Figure 1), and the signal persisted in all sensitivity analyses (Supplementary Figure S2). Of these 47 cases, 20 were classified as infectious, 13 as immune-mediated and 14 as indeterminate (Table 1). Infectious uveitis, chiefly cytomegalovirus chorioretinitis (18 of 20), arose late (median 92 days, range 2–640), was co-reported with another infection in half of cases and had the most serious outcomes (hospitalisation in 10, disability in three and death in two); it was disproportionately reported for both agents (ROR 16.77 and 44.90) and for CD3 bispecific antibodies used outside myeloma (5.76, 3.09–10.74). Immune-mediated uveitis arose early (median 4 days, range 1–11), was co-reported with cytokine release syndrome in 6 of 13 cases and showed a signal only for elranatamab (8 cases; ROR 3.59, 1.79–7.18; IC025 0.42), not for teclistamab (5 cases; 1.26, 0.52–3.03), belantamab mafodotin (0.65, 0.16–2.62), talquetamab (0.89, 0.22–3.58) or CD3 bispecific antibodies used outside myeloma (0.55, 0.28–1.11), although talquetamab showed borderline disproportionality for all uveitis (2.42, 1.08–5.39).

**Table 1.**
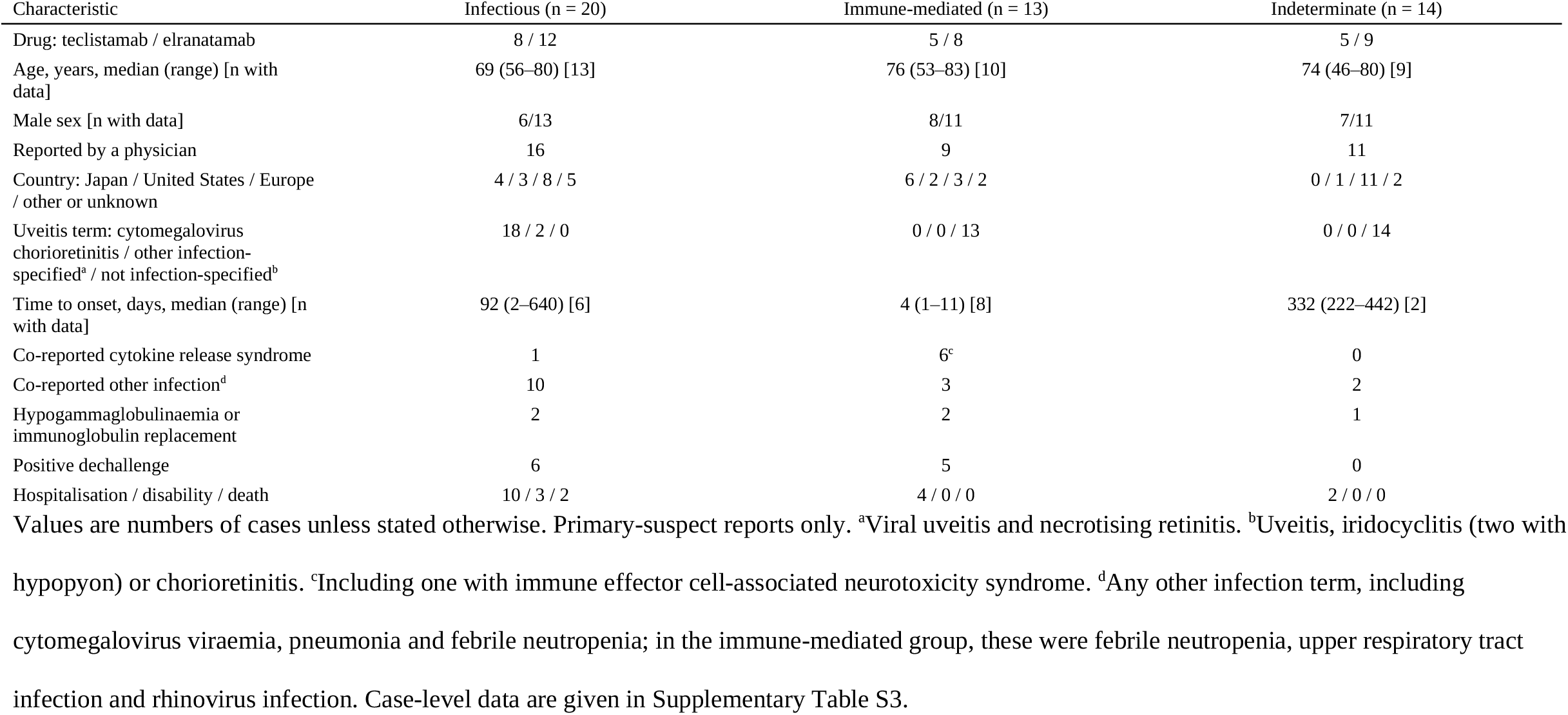
Characteristics of uveitis cases reported with teclistamab or elranatamab, by case classification.

**Figure 1.**
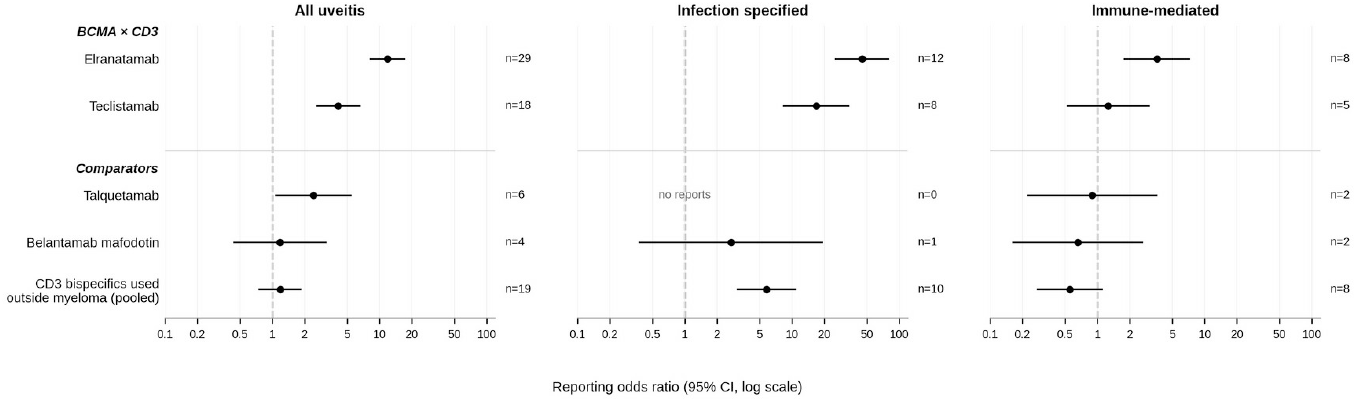
Reporting odds ratios for uveitis with teclistamab, elranatamab and comparator drugs, by outcome level. Points show reporting odds ratios (RORs) and horizontal lines 95% confidence intervals on a logarithmic scale; n, number of uveitis reports. Primary-suspect reports for each drug were compared with all other reports in the database. Left, all uveitis preferred terms (main analysis); middle, reports with an infection-specified term; right, cases classified at case review as immune-mediated (for the reference group, all reports in which no infection was specified were counted). CD3 bispecific antibodies used outside myeloma: mosunetuzumab, glofitamab, epcoritamab, blinatumomab and tarlatamab (pooled).

## Discussion

The FDA uveitis signal therefore appears to comprise two entities: a late infectious uveitis shared by most T-cell–redirecting bispecific antibodies examined and consistent with their profound immunosuppression,^9,10^ and an early immune-mediated uveitis that showed a signal only with elranatamab. With elranatamab, this uveitis arose within days of the first dose (median 2 days, range 1–11), was co-reported with cytokine release syndrome in 4 of 8 cases and abated after drug withdrawal in 3. The lack of a signal with teclistamab, which shares the same target and mechanism, and with belantamab mafodotin, talquetamab and other CD3 bispecific antibodies suggests that neither BCMA targeting nor T-cell redirection per se explains it. Why this entity was confined to elranatamab is uncertain. The two agents differ in antibody structure and dosing: both are given subcutaneously with step-up dosing, but elranatamab at fixed doses and teclistamab at weight-based doses,^1,2^ and early ocular inflammation coinciding with cytokine release syndrome may depend on the magnitude of initial T-cell activation. Reporting patterns may also contribute. A quarter of all elranatamab reports, but only 1% of teclistamab reports, originated from Japan, as did six of the eight elranatamab immune-mediated cases; all-case post-marketing surveillance, often required for newly approved anticancer drugs in Japan, may capture early events more completely, although a population-specific susceptibility cannot be excluded.

Published cases fit this picture. Bilateral cytomegalovirus retinitis has been reported nine months after starting elranatamab,^11^ in keeping with the late infectious entity and with the frequent cytomegalovirus reactivation seen with teclistamab.^10^ A previous disproportionality study of drug-induced immune-mediated uveitis listed both agents among drugs with reporting signals but did not review individual reports;^12^ our case-level review suggests that such screens can conflate infectious and immune-mediated uveitis.

This study has limitations. Spontaneous reports are subject to under-reporting, stimulated reporting after publications and regulatory communications, and incomplete information: onset dates were missing in most reports, ophthalmological details such as anatomical location and aqueous humour testing were rarely available, and classification therefore relied on reported terms, leaving 14 cases indeterminate. The immune-mediated signal rested on eight cases, most from one country. Disproportionality analyses cannot establish causation or incidence; these findings are therefore hypothesis-generating and warrant confirmation in clinical cohorts with systematic ophthalmological assessment. Nonetheless, new visual symptoms in patients receiving these agents warrant prompt ophthalmological assessment, and cytomegalovirus retinitis should be excluded before corticosteroids are started.

## Data Availability

All data used in this study are publicly available from the US Food and Drug Administration Adverse Event Reporting System (FAERS) quarterly data files (https://fis.fda.gov/extensions/FPD-QDE-FAERS/FPD-QDE-FAERS.html). Case-level data for all reviewed reports are provided in the Supplementary Information. The analysis code is available from the corresponding author on reasonable request.

## Acknowledgements

Funding: The authors received no specific funding for this work. Author contributions: Y.W. performed data curation, formal analysis and case classification, and wrote the original draft.

Y.G. conceived the study, provided supervision and clinical interpretation, reviewed the case classifications, and reviewed and edited the manuscript. Both authors approved the final version of the manuscript. Y.W. and Y.G. contributed equally to this work and share first authorship. Artificial intelligence assistance: Claude (Anthropic) was used to assist with writing data-processing and statistical code and with drafting the first version of the manuscript. All code, analyses, case classifications and text were reviewed, verified and edited by the authors, who take full responsibility for the content of this article.

## Data availability statement

The data that support the findings of this study are publicly available from the US Food and Drug Administration Adverse Event Reporting System quarterly data files (https://fis.fda.gov/extensions/FPD-QDE-FAERS/FPD-QDE-FAERS.html). The analysis code is available from the corresponding author on reasonable request.

## Ethics approval statement

This study used publicly available, de-identified data and was therefore exempt from institutional review board approval.

## Patient consent statement

Not applicable.

## Permission to reproduce material from other sources

Not applicable.

## Clinical trial registration

Not applicable.

## References

1. Moreau P, Garfall AL, van de Donk N878J, Nahi H, San-Miguel JF, Oriol A, et al. Teclistamab in relapsed or refractory multiple myeloma. N Engl J Med. 2022;387(6):495–505. doi:10.1056/NEJMoa2203478

2. Lesokhin AM, Tomasson MH, Arnulf B, Bahlis NJ, Miles Prince H, Niesvizky R, et al. Elranatamab in relapsed or refractory multiple myeloma: phase 2 MagnetisMM-3 trial results. Nat Med. 2023;29(9):2259–2267. doi:10.1038/s41591-023-02528-9

3. Castilho Barbosa G, Raza S, Chowdhary A, Sharma S. Manifestations of teclistamab-associated uveitis. EJHaem. 2025;6(3):e70083. doi:10.1002/jha2.70083

4. US Food and Drug Administration. January–March 2026: new safety information or potential signals of serious risks identified by the FDA Adverse Event Monitoring System (AEMS). Updated June 30, 2026. Accessed September 26, 2026. https://www.fda.gov/drugs/fda-adverse-event-monitoring-system-aems/january-march-2026-new-safety-information-or-potential-signals-serious-risks-identified-fda-adverse

5. van Puijenbroek EP, Bate A, Leufkens HG, Lindquist M, Orre R, Egberts AC. A comparison of measures of disproportionality for signal detection in spontaneous reporting systems for adverse drug reactions. Pharmacoepidemiol Drug Saf. 2002;11(1):3–10. doi:10.1002/pds.668

6. Norén GN, Hopstadius J, Bate A. Shrinkage observed-to-expected ratios for robust and transparent large-scale pattern discovery. Stat Methods Med Res. 2013;22(1):57–69. doi:10.1177/0962280211403604

7. Chari A, Minnema MC, Berdeja JG, Oriol A, van de Donk N878J, Rodríguez-Otero P, et al. Talquetamab, a T-cell-redirecting GPRC5D bispecific antibody for multiple myeloma. N Engl J Med. 2022;387(24):2232–2244. doi:10.1056/NEJMoa2204591

8. Fusaroli M, Salvo F, Begaud B, AlShammari TM, Bate A, Battini V, et al. The Reporting of a Disproportionality Analysis for Drug Safety Signal Detection Using Individual Case Safety Reports in PharmacoVigilance (READUS-PV): development and statement. Drug Saf. 2024;47(6):575–584. doi:10.1007/s40264-024-01421-9

9. Mazahreh F, Mazahreh L, Schinke C, Thanendrarajan S, Zangari M, Shaughnessy JD, et al. Risk of infections associated with the use of bispecific antibodies in multiple myeloma: a pooled analysis. Blood Adv. 2023;7(13):3069–3074. doi:10.1182/bloodadvances.2022009435

10. Cheema H, Shrestha A, Naqvi S, Muttineni S, Dronavalli S, Alrawabdeh J, etal. Clinical outcomes and risk factors of cytomegalovirus reactivation in teclistamab-treated multiple myeloma patients. Blood Cancer J. 2026;16(1):51. doi:10.1038/s41408-026-01484-0

11. Cochran-Perez MR, Sharma P, Delisle JS, Schlesinger M, Rao H, Trias J, *etal*. Bispecific antibody therapy-associated opportunistic infections: cytomegalovirus retinitis and chronic salmonellosis in a non-HIV patient. Haematologica.2025;110(11):2855–2858. doi:10.3324/haematol.2025.287351

12. Xu S, Song Z, Li Y, Bai J, Wang D, Wang E, et al. Drug-induced immune-mediated uveitis: a real-world pharmacovigilance analysis using FAERS and JADER databases. BMJ Open Ophthalmol. 2025;10(1):e002456. doi:10.1136/bmjophth-2025-002456

