## Supplementary Information for "Uveitis with teclistamab and elranatamab: infection or inflammation?"

##### Contents

Supplementary Figure S1. Flow diagram of report selection

Supplementary Table S1. Outcome definitions

Supplementary Table S2. Disproportionality results by outcome level

Supplementary Figure S2. Sensitivity analyses

Supplementary Table S3. Case-level listing of reviewed reports

### Supplementary Figure S1. Flow diagram of report selection

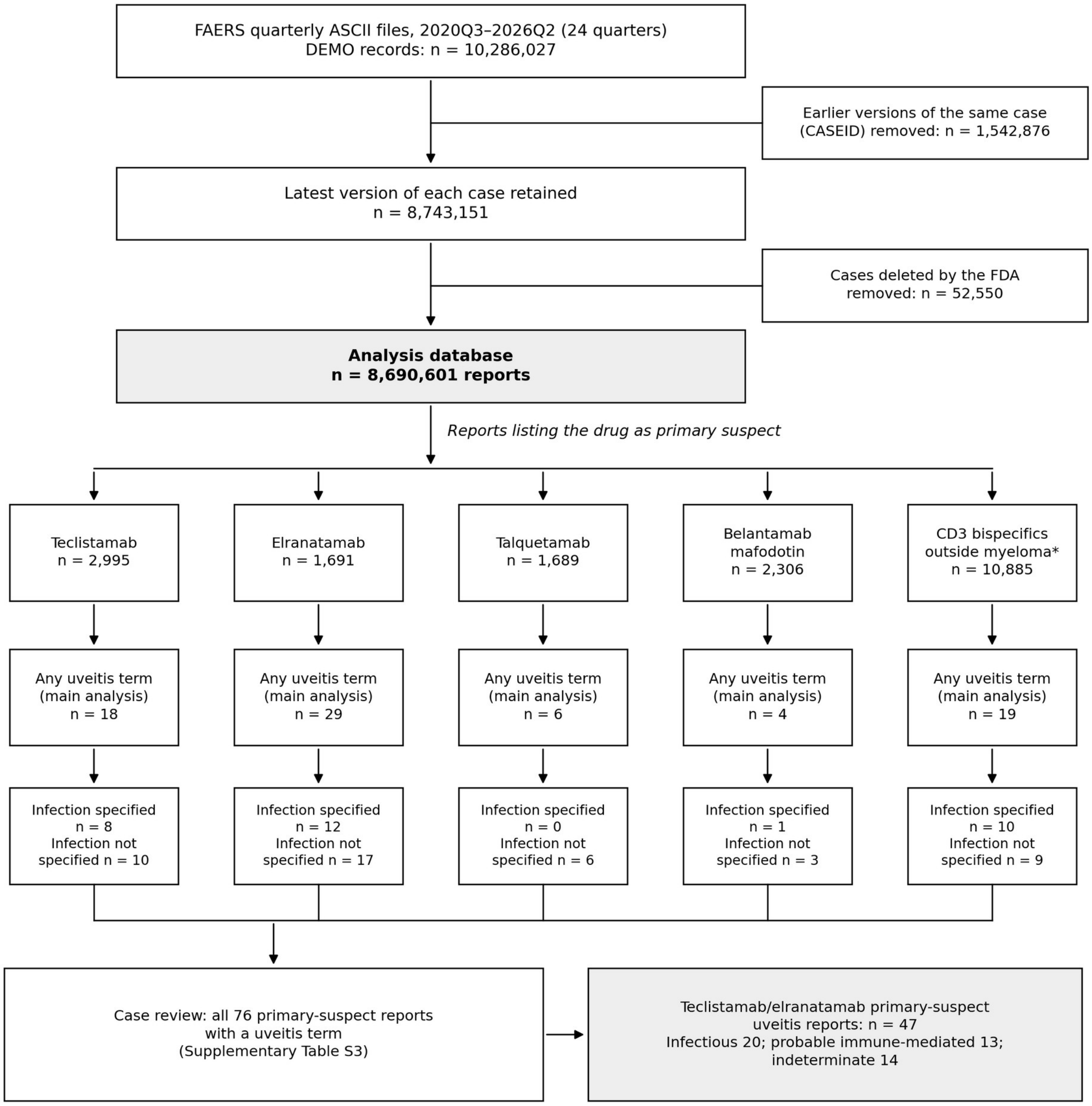


*Numbers of primary-suspect reports and uveitis reports correspond to Supplementary Table S2. *Mosunetuzumab, glofitamab and epcoritamab (CD20 × CD3), blinatumomab (CD19 × CD3) and tarlatamab (DLL3 × CD3), pooled.*

#### Supplementary Table S1. Outcome definitions (MedDRA preferred terms)

FAERS quarterly ASCII files (2020Q3–2026Q2) were downloaded on 25 September 2026 and deduplicated as described in Supplementary Figure S1. Drugs were matched in the DRUGNAME and PROD_AI fields by generic name, brand name and development code (teclistamab, JNJ-64007957; elranatamab, PF-06863135). CD3 bispecific antibodies used outside myeloma: mosunetuzumab, glofitamab, epcoritamab, odronextamab (no reports), blinatumomab and tarlatamab. Multiple myeloma reports: indication of plasma cell myeloma, multiple myeloma, plasmacytoma, plasma cell leukaemia, myeloma cast nephropathy or myelomatosis, or any report listing a myeloma-specific drug. Analyses were performed in R 4.3.3 (data.table, ggplot2).

| Uveitis, infection specified (n = 18) | Uveitis, infection not specified (n = 19) |
|---|---|
| Cytomegalovirus chorioretinitis | Uveitis |
| Cytomegalovirus retinitis | Iritis |
| Viral uveitis | Iridocyclitis |
| Infective uveitis | Cyclitis |
| Infective iritis | Chorioretinitis |
| Infectious iridocyclitis | Choroiditis |
| Bacterial iritis | Noninfective chorioretinitis |
| Viral keratouveitis | Vitritis |
| Retinitis viral | Panuveitis |
| Necrotising retinitis | Pars planitis |
| Acute retinal necrosis | Immune-mediated uveitis |
| Vitritis infective | Autoimmune uveitis |
| Fungal retinitis | Keratouveitis |
| Candida retinitis | Vogt-Koyanagi-Harada disease |
| Retinitis histoplasma | Serpiginous choroiditis |
| Ocular toxoplasmosis | Birdshot chorioretinopathy |
| Toxoplasma chorioretinitis | Sympathetic ophthalmia |
| Immune recovery uveitis | Tubulointerstitial nephritis and uveitis syndrome |
| | Enteropathic uveitis |

*All uveitis (main analysis) = either column. Probable immune-mediated cases were identified at case review (Supplementary Table S3) among reports in which no infection was specified. Positive control (keratopathy): PTs containing 'keratopathy', 'corneal epithelium', 'corneal disorder', 'corneal deposits', 'corneal cyst' or 'microcyst', PTs ending in 'keratitis', and punctate keratitis.*

#### Supplementary Table S2. Disproportionality results by outcome level (primary-suspect reports vs all other reports)

| Drug (primary-suspect reports) | Outcome | Cases | ROR (95% CI) | IC025 |
|---|---|---|---|---|
| Teclistamab (n = 2,995) | All uveitis (main analysis) | 18 | 4.10 (2.58–6.52) | 1.12 |
| | Infection specified | 8 | 16.77 (8.36–33.64) | 1.90 |
| | Infection not specified | 10 | 2.53 (1.36–4.70) | 0.15 |
| | Probable immune-mediated | 5 | 1.26 (0.52–3.03) | −1.26 |
| Elranatamab (n = 1,691) | All uveitis (main analysis) | 29 | 11.85 (8.21–17.12) | 2.68 |
| | Infection specified | 12 | 44.90 (25.38–79.41) | 3.04 |
| | Infection not specified | 17 | 7.66 (4.75–12.36) | 1.86 |
| | Probable immune-mediated | 8 | 3.59 (1.79–7.18) | 0.42 |
| Talquetamab (n = 1,689) | All uveitis (main analysis) | 6 | 2.42 (1.08–5.39) | −0.29 |
| | Infection specified | 0 | NE | −10.95 |
| | Infection not specified | 6 | 2.69 (1.21–5.99) | −0.17 |
| | Probable immune-mediated | 2 | 0.89 (0.22–3.58) | −2.72 |
| Belantamab mafodotin (n = 2,306) | All uveitis (main analysis) | 4 | 1.18 (0.44–3.14) | −1.56 |
| | Infection specified | 1 | 2.70 (0.38–19.21) | −3.00 |
| | Infection not specified | 3 | 0.98 (0.32–3.05) | −2.09 |
| | Probable immune-mediated | 2 | 0.65 (0.16–2.62) | −3.10 |
| | Keratopathy (positive control) | 930 | 1422.88 (1302.21–1554.74) | 8.87 |
| CD3 bispecific antibodies used outside myeloma (pooled) (n = 10,885) | All uveitis (main analysis) | 19 | 1.19 (0.76–1.86) | −0.53 |
| | Infection specified | 10 | 5.76 (3.09–10.74) | 1.15 |
| | Infection not specified | 9 | 0.62 (0.32–1.20) | −1.79 |
| | Probable immune-mediated | 8 | 0.55 (0.28–1.11) | −2.03 |
| Mosunetuzumab (n = 657) | All uveitis (main analysis) | 5 | 5.20 (2.16–12.54) | 0.34 |
| | Infection specified | 1 | 9.50 (1.34–67.60) | −2.47 |
| | Infection not specified | 4 | 4.62 (1.73–12.34) | −0.05 |
| | Probable immune-mediated | 4 | 4.62 (1.73–12.34) | −0.05 |
| Glofitamab (n = 1,444) | All uveitis (main analysis) | 4 | 1.88 (0.71–5.03) | −0.99 |
| | Infection specified | 3 | 12.99 (4.18–40.38) | 0.19 |
| | Infection not specified | 1 | 0.52 (0.07–3.71) | −4.47 |

| | | | | |
|---|---|---|---|---|
| | Probable immune-mediated | 0 | NE | −12.60 |
| Epcoritamab (n = 1,817) | All uveitis (main analysis) | 6 | 2.25 (1.01–5.01) | −0.38 |
| | Infection specified | 4 | 13.78 (5.16–36.81) | 0.74 |
| | Infection not specified | 2 | 0.83 (0.21–3.32) | −2.81 |
| | Probable immune-mediated | 2 | 0.83 (0.21–3.32) | −2.81 |
| Blinatumomab (n = 5,225) | All uveitis (main analysis) | 3 | 0.39 (0.13–1.21) | −3.30 |
| | Infection specified | 2 | 2.39 (0.60–9.56) | −1.69 |
| | Infection not specified | 1 | 0.14 (0.02–1.02) | −6.09 |
| | Probable immune-mediated | 1 | 0.14 (0.02–1.02) | −6.09 |
| Tarlatamab (n = 1,742) | All uveitis (main analysis) | 1 | 0.39 (0.05–2.77) | −4.81 |
| | Infection specified | 0 | NE | −10.96 |
| | Infection not specified | 1 | 0.43 (0.06–3.07) | −4.69 |
| | Probable immune-mediated | 1 | 0.43 (0.06–3.07) | −4.69 |

*CD3 bispecific antibodies used outside myeloma (pooled): mosunetuzumab, glofitamab and epcoritamab (CD20 × CD3; lymphoma), blinatumomab (CD19 × CD3; acute lymphoblastic leukaemia) and tarlatamab (DLL3 × CD3; small-cell lung cancer); odronextamab had no reports. Talquetamab, the only other CD3 bispecific antibody used in myeloma, is shown separately; with talquetamab included in the pooled group (25 uveitis reports among 12,574), RORs were 1.35 (0.91–2.00) for all uveitis, 4.99 (2.68–9.29) for infection specified, 0.90 (0.54–1.49) for infection not specified and 0.60 (0.32–1.11) for probable immune-mediated uveitis. Each drug compared with all other reports in the database (8,690,601). Signal of disproportionate reporting: lower 95% confidence limit of the ROR >1 with ≥3 cases; confirmed by IC025 >0. Probable immune-mediated cases are a subset of reports in which no infection was specified (reference group counted accordingly). IC025, lower limit of the 95% credibility interval of the information component; NE, not estimable (no reports); ROR, reporting odds ratio.*

#### Supplementary Figure S2. Sensitivity analyses (all uveitis)

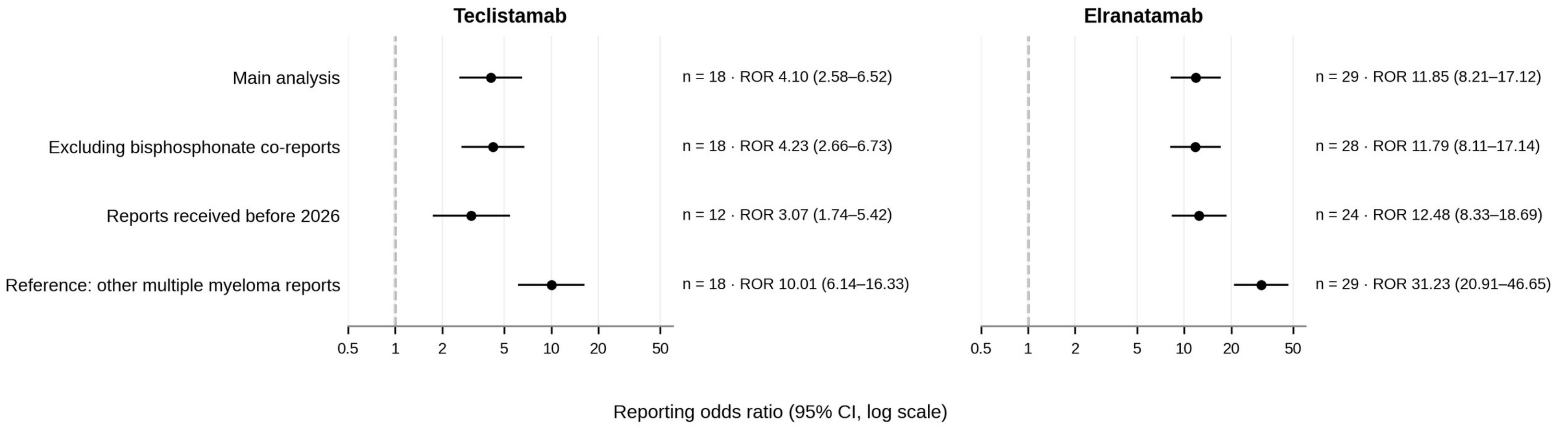


*Reporting odds ratios (ROR) with 95% confidence intervals on a logarithmic scale; n, number of uveitis reports. RORs are larger with the multiple myeloma reference group because uveitis is rarely reported among multiple myeloma reports.*

#### Supplementary Table S3. Case-level listing of reviewed reports

| Case | Drug (role) | Uveitis-related preferred term(s) | Final classification | Basis for classification | Time to onset (days) | Dechallenge | Age/sex | Reporter (country) | Serious outcome(s) |
|---|---|---|---|---|---|---|---|---|---|
| 1 | Teclistamab(PS) | Cytomegalovirus chorioretinitis | Infectious | Cytomegalovirus chorioretinitis term | Not reported | Unknown | 70/NR | Physician (CH) | Other serious |
| 2 | Teclistamab(PS); Teclistamab(SS) | Cytomegalovirus chorioretinitis | Infectious | Cytomegalovirus chorioretinitis term | 31 | Positive | 64/F | Physician (FR) | Hospitalisation; Other serious |
| 3 | Teclistamab(PS) | Cytomegalovirus chorioretinitis | Infectious | Cytomegalovirus chorioretinitis term | 91 | Negative | 71/M | Physician (BE) | Disability; Hospitalisation; Other serious |
| 4 | Teclistamab(PS) | Necrotising retinitis | Infectious | Necrotising retinitis term (presumed infectious; confirmed by second reviewer) | 92 | Not reported | 70/M | Pharmacist (FR) | Hospitalisation |
| 5 | Teclistamab(PS) | Cytomegalovirus chorioretinitis | Infectious | Cytomegalovirus chorioretinitis term | Not reported | Unknown | NR/NR | Physician (US) | Other serious |
| 6 | Teclistamab(PS) | Cytomegalovirus chorioretinitis | Infectious | Cytomegalovirus chorioretinitis term | Not reported | Unknown | NR/NR | Physician (BR) | Other serious |
| 7 | Teclistamab(PS) | Cytomegalovirus chorioretinitis | Infectious | Cytomegalovirus chorioretinitis term | Not reported | Negative | 77/M | NR (EU) | Disability |
| 8 | Teclistamab(PS) | Cytomegalovirus chorioretinitis | Infectious | Cytomegalovirus chorioretinitis term | Not reported | Not reported | NR/F | Physician (US) | Other serious |
| 9 | Teclistamab(PS) | Hypopyon; Iridocyclitis | Probable immune-mediated | No ocular pathogen; onset ≤30 days, positive dechallenge | 7 | Positive | 62/F | Physician (NR) | Required intervention |
| 10 | Teclistamab(PS) | Uveitis | Probable immune-mediated | No ocular pathogen; CRS/ICANS co-reported | Not reported | Unknown | NR/NR | Physician (US) | Other serious |
| 11 | Teclistamab(PS) | Uveitis | Probable immune-mediated | No ocular pathogen; CRS/ICANS co-reported | Not reported | Unknown | NR/M | Physician (FR) | Other serious |
| 12 | Teclistamab(PS) | Uveitis | Probable immune-mediated | No ocular pathogen; positive dechallenge | Not reported | Positive | 83/F | Other HP (US) | Other serious |
| 13 | Teclistamab(PS) | Iridocyclitis | Probable immune-mediated | No ocular pathogen; onset ≤30 days | 5 | Not reported | 76/M | Pharmacist (EU) | Hospitalisation |
| 14 | Teclistamab(PS) | Chorioretinitis | Indeterminate | No ocular pathogen and no immune-mediated criterion | Not reported | Not reported | NR/M | Physician (BR) | Other serious |
| 15 | Teclistamab(PS) | Uveitis | Indeterminate | No ocular pathogen and no immune-mediated criterion | Not reported | Not reported | 76/M | Other HP (US) | Other serious |
| 16 | Teclistamab(PS) | Uveitis | Indeterminate | No ocular pathogen and no immune-mediated criterion | Not reported | Not reported | NR/NR | Physician (EU) | Other serious |
| 17 | Teclistamab(PS) | Uveitis | Indeterminate | No ocular pathogen and no immune-mediated criterion | Not reported | Negative | 80/F | Physician (EU) | Other serious |
| 18 | Teclistamab(PS) | Uveitis | Indeterminate | No ocular pathogen and no immune-mediated criterion | 442 | Negative | 46/M | NR (EU) | Other serious |

| | | | | | | | | | |
|---|---|---|---|---|---|---|---|---|---|
| 19 | Elranatamab(PS) | Cytomegalovirus chorioretinitis | Infectious | Cytomegalovirus chorioretinitis term | Not reported | Unknown | NR/NR | Other HP (GB) | Other serious |
| 20 | Elranatamab(PS); Elranatamab(SS) | Cytomegalovirus chorioretinitis | Infectious | Cytomegalovirus chorioretinitis term; anti-cytomegalovirus therapy co-reported | Inconsistent dates | Positive | 69/M | Physician (JP) | Hospitalisation; Other serious |
| 21 | Elranatamab(PS) | Cytomegalovirus chorioretinitis | Infectious | Cytomegalovirus chorioretinitis term | 267 | Positive | 64/F | Physician (BE) | Disability; Hospitalisation; Other serious |
| 22 | Elranatamab(PS) | Cytomegalovirus chorioretinitis | Infectious | Cytomegalovirus chorioretinitis term | Not reported | Unknown | NR/NR | Physician (TW) | Other serious |
| 23 | Elranatamab(PS); Elranatamab(SS) | Cytomegalovirus chorioretinitis | Infectious | Cytomegalovirus chorioretinitis term | 640 | Positive | 56/F | Physician (TR) | Death; Hospitalisation; Other serious |
| 24 | Elranatamab(PS) | Cytomegalovirus chorioretinitis | Infectious | Cytomegalovirus chorioretinitis term | Not reported | Unknown | 64/M | Physician (TR) | Other serious |
| 25 | Elranatamab(PS); Elranatamab(SS) | Cytomegalovirus chorioretinitis | Infectious | Cytomegalovirus chorioretinitis term | Not reported | Negative | NR/NR | Pharmacist (JP) | Other serious |
| 26 | Elranatamab(PS); Elranatamab(SS) | Cytomegalovirus chorioretinitis | Infectious | Cytomegalovirus chorioretinitis term | Not reported | Unknown | 75/M | Physician (EU) | Death; Other serious |
| 27 | Elranatamab(PS) | Cytomegalovirus chorioretinitis | Infectious | Cytomegalovirus chorioretinitis term | Not reported | Positive | NR/NR | Physician (JP) | Hospitalisation; Life-threatening; Other serious |
| 28 | Elranatamab(PS) | Cytomegalovirus chorioretinitis | Infectious | Cytomegalovirus chorioretinitis term | Not reported | Not applicable | 80/F | Physician (US) | Hospitalisation; Other serious |
| 29 | Elranatamab(PS) | Cytomegalovirus chorioretinitis | Infectious | Cytomegalovirus chorioretinitis term; early onset, possibly pre-existing infection | 2 | Positive | 62/F | Physician (JP) | Hospitalisation |
| 30 | Elranatamab(PS) | Viral uveitis | Infectious | Viral uveitis term | Not reported | Unknown | 68/F | Physician (IL) | Hospitalisation; Other serious |
| 31 | Elranatamab(PS) | Uveitis | Probable immune-mediated | No ocular pathogen; onset ≤30 days | 11 | Not reported | NR/M | Consumer (BR) | Hospitalisation; Other serious |
| 32 | Elranatamab(PS); Elranatamab(SS) | Uveitis | Probable immune-mediated | No ocular pathogen; onset ≤30 days, positive dechallenge | 1 | Positive | 76/F | Physician (JP) | Hospitalisation |
| 33 | Elranatamab(PS) | Uveitis | Probable immune-mediated | No ocular pathogen; onset ≤30 days, CRS/ICANS co-reported | 1 | Unknown | 72/M | Physician (JP) | Other serious |
| 34 | Elranatamab(PS) | Uveitis | Probable immune-mediated | No ocular pathogen; CRS/ICANS co-reported | Not reported | Unknown | 69/M | Physician (IT) | Hospitalisation |
| 35 | Elranatamab(PS); Elranatamab(SS) | Uveitis | Probable immune-mediated | No ocular pathogen; CRS/ICANS co-reported, positive dechallenge | Inconsistent dates | Positive | 53/M | Physician (JP) | Other serious |
| 36 | Elranatamab(PS) | Uveitis | Probable immune-mediated | No ocular pathogen; onset ≤30 days | 3 | Unknown | 80/NR | Physician (JP) | Other serious |
| 37 | Elranatamab(PS) | Uveitis | Probable immune-mediated | No ocular pathogen; onset ≤30 days | 5 | Not reported | 76/M | Physician (JP) | Other serious |
| 38 | Elranatamab(PS) | Hypopyon; Uveitis | Probable immune-mediated | No ocular pathogen; onset ≤30 days, CRS/ICANS co-reported, positive dechallenge | 1 | Positive | 81/M | Pharmacist (JP) | Other serious |
| 39 | Elranatamab(PS) | Uveitis | Indeterminate | No ocular pathogen and no immune-mediated criterion | Not reported | Negative | 69/F | Physician (FR) | Hospitalisation |
| 40 | Elranatamab(PS) | Uveitis | Indeterminate | No ocular pathogen and no immune-mediated criterion; bisphosphonate co-reported (causal role unclear) | 222 | Negative | 76/F | Physician (FR) | Hospitalisation |
| 41 | Elranatamab(PS); Elranatamab(SS) | Uveitis | Indeterminate | No ocular pathogen and no immune-mediated criterion | Not reported | Not reported | 76/M | Physician (FR) | Other serious |
| 42 | Elranatamab(PS) | Uveitis | Indeterminate | No ocular pathogen and no immune-mediated criterion | Not reported | Unknown | NR/NR | Physician (FR) | Other serious |
| 43 | Elranatamab(PS) | Uveitis | Indeterminate | No ocular pathogen and no immune-mediated criterion | Not reported | Unknown | NR/NR | Physician (FR) | Other serious |
| 44 | Elranatamab(PS) | Uveitis | Indeterminate | No ocular pathogen and no immune-mediated criterion | Not reported | Unknown | NR/M | Pharmacist (GB) | Other serious |
| 45 | Elranatamab(PS) | Uveitis | Indeterminate | No ocular pathogen and no immune-mediated criterion | Not reported | Not reported | 69/M | Physician (EU) | Other serious |
| 46 | Elranatamab(PS) | Uveitis | Indeterminate | No ocular pathogen and no immune-mediated criterion | Not reported | Not reported | 74/F | Physician (EU) | Other serious |
| 47 | Elranatamab(PS) | Iridocyclitis | Indeterminate | Non-ocular infections co-reported without an ocular pathogen; no immune-mediated criterion | Not reported | Not reported | 68/M | Physician (IL) | Other serious |
| 48 | Talquetamab(PS); Talquetamab(SS) | Uveitis | Probable immune-mediated | No ocular pathogen; positive dechallenge | Not reported | Positive | NR/F | Physician (FI) | Other serious |
| 49 | Talquetamab(PS) | Uveitis | Probable immune-mediated | No ocular pathogen; positive dechallenge | Not reported | Positive | NR/NR | Physician (JP) | Other serious |
| 50 | Talquetamab(PS) | Iritis | Indeterminate | No ocular pathogen and no immune-mediated criterion | Not reported | Unknown | NR/NR | Physician (US) | Not reported |
| 51 | Talquetamab(PS) | Uveitis | Indeterminate | No ocular pathogen and no immune-mediated criterion | Not reported | Unknown | NR/NR | Physician (BR) | Other serious |
| 52 | Talquetamab(PS) | Uveitis | Indeterminate | No ocular pathogen and no immune-mediated criterion | Not reported | Unknown | NR/NR | Physician (HK) | Other serious |
| 53 | Talquetamab(PS) | Uveitis | Indeterminate | No ocular pathogen and no immune-mediated criterion | Not reported | Unknown | NR/NR | Physician (US) | Other serious |
| 54 | Belantamab(PS) | Cytomegalovirus chorioretinitis | Infectious | Cytomegalovirus chorioretinitis term | 114 | Positive | 64/F | Physician (IL) | Hospitalisation; Other serious |
| 55 | Belantamab(PS) | Iritis | Probable immune-mediated | No ocular pathogen; positive dechallenge | Not reported | Positive | 70/M | Physician (US) | Other serious |
| 56 | Belantamab(PS) | Iritis | Probable immune-mediated | No ocular pathogen; positive dechallenge | Not reported | Positive | 61/M | Other HP (US) | Other serious |

| | | | | | | | | | |
|---|---|---|---|---|---|---|---|---|---|
| 57 | Belantamab(PS) | Uveitis | Indeterminate | No ocular pathogen and no immune-mediated criterion | 56 | Not reported | NR/NR | Physician (US) | Other serious |
| 58 | Mosunetuzumab(PS) | Necrotising retinitis; Retinitis | Infectious | Necrotising retinitis term (presumed infectious) | 159 | Positive | 76/M | Physician (JP) | Hospitalisation; Other serious |
| 59 | Mosunetuzumab(PS) | Uveitis | Probable immune-mediated | No ocular pathogen; onset ≤30 days, positive dechallenge | 9 | Positive | 73/M | Physician (NR) | Disability; Hospitalisation |
| 60 | Mosunetuzumab(PS) | Uveitis | Probable immune-mediated | No ocular pathogen; onset ≤30 days, positive dechallenge | 2 | Positive | 59/M | Physician (NR) | Hospitalisation |
| 61 | Mosunetuzumab(PS) | Uveitis | Probable immune-mediated | No ocular pathogen; onset ≤30 days, positive dechallenge | 21 | Positive | 37/M | Physician (NR) | Other serious |
| 62 | Mosunetuzumab(PS) | Iridocyclitis | Probable immune-mediated | No ocular pathogen; positive dechallenge | Not reported | Positive | 55/M | Physician (EU) | Other serious |
| 63 | Glofitamab(PS) | Necrotising retinitis | Infectious | Necrotising retinitis term (presumed infectious); anti-cytomegalovirus therapy co-reported | 86 | Not reported | 73/M | Physician (NR) | Hospitalisation |
| 64 | Glofitamab(PS) | Cytomegalovirus chorioretinitis | Infectious | Cytomegalovirus chorioretinitis term | Not reported | Unknown | NR/NR | Physician (NR) | Other serious |
| 65 | Glofitamab(PS) | Cytomegalovirus chorioretinitis | Infectious | Cytomegalovirus chorioretinitis term | 235 | Positive | 51/M | NR (CN) | Hospitalisation |
| 66 | Glofitamab(PS) | Uveitis | Indeterminate | No ocular pathogen and no immune-mediated criterion | Not reported | Unknown | NR/F | Pharmacist (US) | Other serious |
| 67 | Epcoritamab(PS) | Cytomegalovirus chorioretinitis | Infectious | Cytomegalovirus chorioretinitis term | Inconsistent dates | Unknown | 80/F | Physician (JP) | Other serious |
| 68 | Epcoritamab(PS) | Cytomegalovirus chorioretinitis | Infectious | Cytomegalovirus chorioretinitis term | Inconsistent dates | Not applicable | 72/M | Physician (JP) | Death; Hospitalisation; Other serious |
| 69 | Epcoritamab(PS) | Cytomegalovirus chorioretinitis | Infectious | Cytomegalovirus chorioretinitis term; early onset, possibly pre-existing infection | 18 | Positive | 74/F | Physician (NR) | Other serious |
| 70 | Epcoritamab(PS) | Cytomegalovirus chorioretinitis | Infectious | Cytomegalovirus chorioretinitis term | Not reported | Not reported | 70/NR | Physician (NR) | Other serious |
| 71 | Epcoritamab(PS) | Uveitis | Probable immune-mediated | No ocular pathogen; positive dechallenge | Not reported | Positive | NR/M | Physician (US) | Other serious |
| 72 | Epcoritamab(PS) | Uveitis | Probable immune-mediated | No ocular pathogen; CRS/ICANS co-reported | Inconsistent dates | Unknown | 80/NR | Physician (JP) | Other serious |
| 73 | Blinatumomab(PS) | Cytomegalovirus chorioretinitis | Infectious | Cytomegalovirus chorioretinitis term; anti-cytomegalovirus therapy co-reported; early onset, possibly pre-existing infection | 0 | Positive | 59/M | Physician (JP) | Disability; Hospitalisation; Other serious |
| 74 | Blinatumomab(PS) | Cytomegalovirus chorioretinitis | Infectious | Cytomegalovirus chorioretinitis term; anti-cytomegalovirus therapy co-reported | Not reported | Not applicable | 2/M | Other HP (US) | Death; Other serious |
| 75 | Blinatumomab(PS) | Iridocyclitis | Probable immune-mediated | No ocular pathogen; CRS/ICANS co-reported | Not reported | Not applicable | NR/NR | Other HP (US) | Death; Hospitalisation; Other serious |
| 76 | Tarlatamab(PS) | Uveitis | Probable immune-mediated | No ocular pathogen; onset ≤30 days | 3 | Not reported | NR/M | Pharmacist (JP) | Other serious |

*All 76 primary-suspect reports with a uveitis term for teclistamab (18), elranatamab (29), talquetamab (6), belantamab mafodotin (4) and CD3 bispecific antibodies used outside myeloma (mosunetuzumab 5, glofitamab 4, epcoritamab 6, blinatumomab 3, tarlatamab 1) were reviewed. Classification criteria: infectious, pathogen-specific ocular term, co-reported pathogen-specific infection or anti-cytomegalovirus therapy; probable immune-mediated, no infectious evidence and ≥1 of onset ≤30 days after the first dose, co-reported cytokine release syndrome (CRS) or immune effector cell-associated neurotoxicity syndrome (ICANS), or positive dechallenge/rechallenge; indeterminate, neither. Classification was performed by a physician blinded to a rule-based algorithm applying the same criteria (agreement 75/76, 98.7%; κ = 0.98); the single discrepancy was resolved in favour of the reviewer's classification, and one uncertain case (necrotising retinitis with teclistamab) was reviewed by a haematologist-oncologist. Possible duplicates: identical age, sex, country and event date. Time to onset: event date minus first-dose date; negative values indicate inconsistent dates. CMV, cytomegalovirus; ICI, immune checkpoint inhibitor; IVIG, intravenous immunoglobulin; NR, not reported; PS, primary suspect; SS, secondary suspect.*
